# Nutrition Able: A Novel Tool for Improving and Assessing Nutritional Literacy of Middle School Children

**DOI:** 10.1101/2025.07.20.25331738

**Authors:** Jeffery Bolte, Jacob Abalos, Jakson Johnson, Alejandra Garcia, Alex Joyce, Christopher Hinds, Paarth Patel, Esther Nnali, Victoria Vargas, Allison Stewart, Avinash Kakulavar, Jeff Svec

**Affiliations:** University of Texas Health Science Center at San Antonio Long School of Medicine; University of Texas at San Antonio

**Author notes:** **Corresponding author:** Jeffery Bolte, 7403 Wurzbach Rd Apt 266 San Antonio TX 78229.

## Abstract

**Background:** Overweight and obesity are leading contributors to chronic disease in Bexar County, with deficiencies in health education playing a central role in their prevalence. Nutrition Able is a medical student–led community initiative from the University of Texas Health Science Center at San Antonio (UTHSCSA) designed to improve nutritional literacy among school-aged children and to pilot a novel assessment instrument. As part of the intervention, the Nutrition Able team delivers classroom-based educational sessions focused on understanding the futrition facts label. To evaluate the impact of this intervention, the Nutrition Able Literacy Assessment (NALA)—a 7-item pre/post survey—was developed to measure functional knowledge of core nutritional concepts. In its pilot, NALA demonstrated consistent performance, aligning with known trends in nutritional literacy literature and offering high-throughput assessment with minimal time burden.

Pilot results revealed that attending school in a food desert was associated with lower baseline NALA scores, and subjects at the extremes of BMI had lower performance, with the highest performance in the overweight/obese categories. Female students scored higher when the session was led by a female presenter, suggesting that educator-student demographic concordance may influence educational outcomes. Post-intervention scores improved across all groups, supporting the effectiveness of both the educational session and the NALA instrument in capturing knowledge gains. In conclusion, Nutrition Able improves youth understanding of nutrition label information, and the NALA tool shows promise as a brief, scalable instrument for evaluating nutrition literacy in public health and clinical settings. These findings also highlight the potential impact of educator demographics on health education outcomes, suggesting that demographically matched educators may enhance intervention efficacy.

## Introduction

Eating patterns are influenced by culture^(1)^, socioeconomic status^(2)^, physiological state^(3)^, psychological state^(4)^, time of day^(5,6)^, geography^7,8^, and nutritional literacy^9^. Barriers to optimizing eating patterns include nutrition/health literacy, geography, finances, and time^10,11^. Physicians cannot address all aspects of the complex biopsychosocial inputs to diet within time-constrained appointments, so they must help patients self-regulate by increasing health literacy. Ratzan and Parker defined health literacy as “The degree to which individuals have the capacity to obtain, process, and understand basic health information and services needed to make appropriate health decisions”^12^. Evidence-based physicians, dietitians and diabetes educators should focus on educating patients about regulation of their food environments with health in mind^12^.

In Texas, ∼2.7 million people (12.3% of the adult population) have diabetes^13^. Estimates suggest that another 621,000 people have diabetes but are undiagnosed^14^. Bexar County has higher diabetes rates than the state average, with over 100,000 people diagnosed (between 13%-15% of the population)^15^. Approximately 72% of the population is overweight, obese, or has pre-diabetes, predisposing the population to high rates of cardiovascular disease, stroke, Alzheimer’s disease, and cancer^15^.

Diet quality is a determinant of health. Diets that reduce the risk of metabolic syndrome feature nutrient-dense foods that are high in fiber and protein and low in saturated fat^(16–20)^. These foods have low potential for overconsumption^21^. Calories drive changes in body fat and risk of metabolic syndrome^16,22^. While many factors that influence eating behaviors and diet quality are functionally nonmodifiable from the medical practitioner’s perspective, nutritional literacy is a cost-efficient modifiable component determining diet quality. This echos research showing that education status is a predictor of health outcomes^23^. For instance, diabetic education is a viable way of improving glycemic control in patients with diabetes^24^, and improving nutritional literacy can help prevent lifelong complications of poor diet quality^9,25^.

### Nutrition Facts Labels’ Importance in Food Deserts

Another determinant of diet quality is geography^8^. The USDA measures the influence of geography on food scarcity by looking at a variety of factors which are classified into varying degrees of a “low-income low access tract”, colloquially known as a food desert (FD). Food scarcity is measured using income level, transportation availability, and distance from major supermarkets. For instance, one type of FD, a “low-income and low-access tract measured at 1 and 10 miles”, is defined as “a low-income tract with at least 500 people, or 33 percent of the population, living more than 1 mile (urban areas) or more than 10 miles (rural areas) from the nearest supermarket, supercenter, or large grocery store”^26^.

Nutrition facts labels are the primary line of communication of nutritional information to the consumer. The FDA mandates that many foods, including all processed foods, contain a nutrition facts label. All foods that make any sort of claim about health implications or nutrient content must display an FDA-compliant nutrition label^27^. Notable exceptions are any whole, unprocessed foods such as raw fruits, vegetables, fish, and meats.

FDs may be more than just barriers to healthy foods-they may be barriers to literacy. Intervening on nutritional literacy at a young age may encourage lifelong, incrementally advantageous food decisions in time and resource-constrained environments. In FDs, children’s meals often come from school-provided lunches, home-grown food, nearby convenience stores, and fast food restaurants^28^. For young students in these FDs, few whole food options are available. Most available foods will have nutrition labels, underscoring the importance of educating children about how and why to use a nutrition label.

### Development and Structure of the NALA Instrument

The Nutrition Able team developed a lecture, How to Read a Nutrition Label, which seeks address this disparity. A novel assessment tool was developed to measure participants’ level of functional understanding of the core components and health relevance of the label: calories; macronutrients; calories per serving; applied macronutrient understanding; fiber; protein; and lifestyle interventions to prevent diabetes and obesity.

The Nutrition Able Literacy Assessment (NALA) (Figure 1) is an original, 7-item multiple-choice and short-response (for demographics) tool designed to assess core components of nutrition label literacy. The assessment includes questions targeting knowledge of calories, macronutrients, serving size calculations, fiber, and protein, as well as their relevance to health outcomes. It can be administered in 5 minutes or less to a large class of students, making it convenient for large scale school based literacy assessments. Demographic items (e.g., age, gender, grade level) and family history of metabolic disease are also collected to allow for subgroup analyses. The tool was reviewed by members of the University of Texas Health Science Center Diabetes Division for face validity.

**Figure 1:**
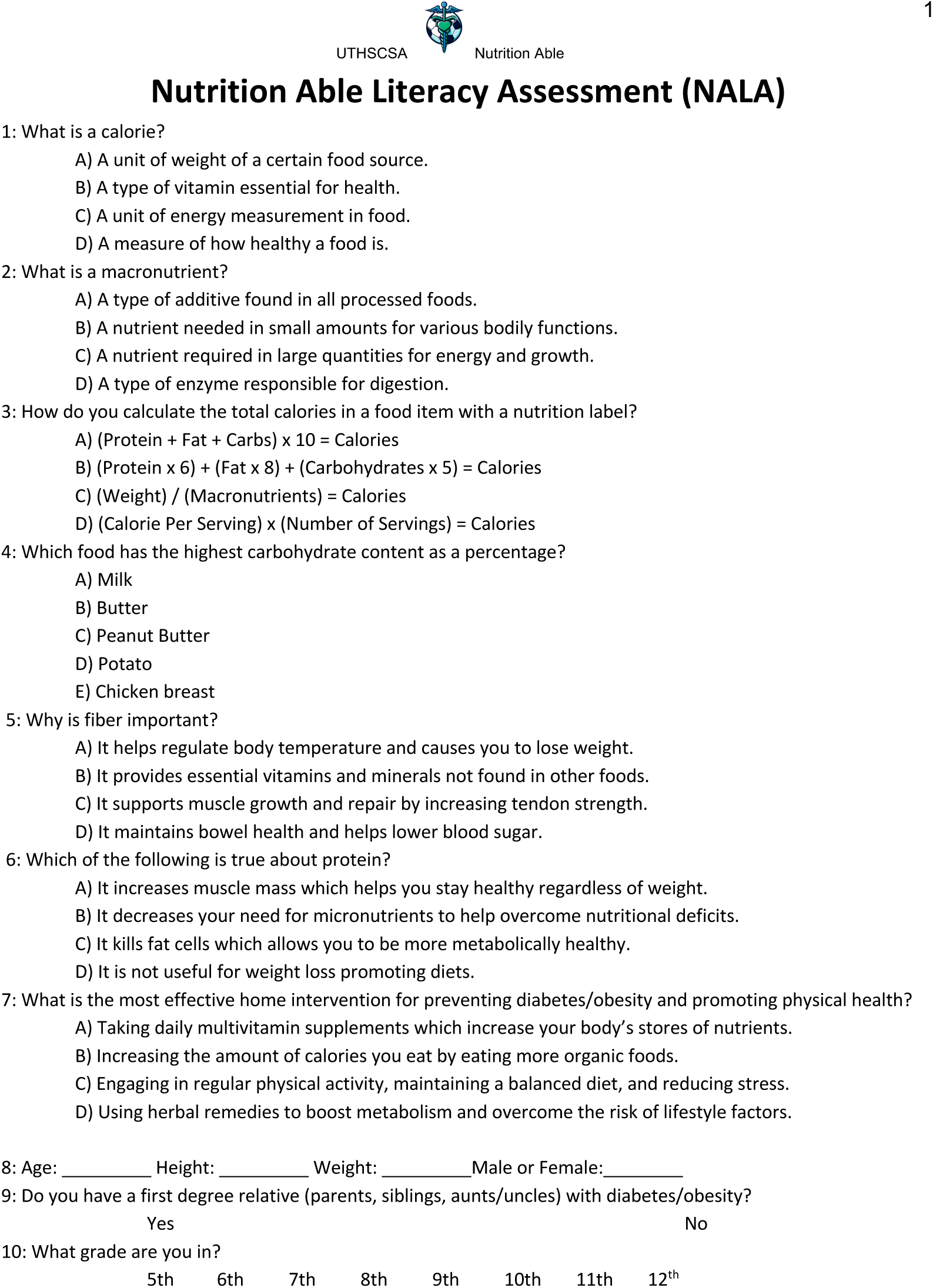

### Pilot Implementation and Study Design

This study is a pilot evaluation of the NALA tool as part of a school-based educational intervention entitled Nutrition Able: How to Read a Nutrition Label. The pilot was conducted with middle and high school students primarily in a local public school district. Participants completed NALA before and after receiving the Nutrition Able lecture. The primary aim of this pilot is to assess the feasibility, clarity, and preliminary utility of the NALA instrument in capturing representative and reproducible data with respect to functional nutrition facts label literacy.

### The Need for a New Tool

Previous studies have emphasized the importance of school-based, early-life nutritional/food literacy interventions^29,30^. Dieticians have lobbied to address disparities in health literacy in FDs for decades^31^, citing well-studied relationships between food insecurity and nutritional literacy^32^. Much of the research in this space assesses either the behavioral components of nutritional literacy or the financial components of food insecurity. Fewer studies directly assess knowledge-based nutritional literacy. Vidgen and Galledos wrote that “‘Food literacy’ has emerged as a term to describe the everyday practicalities associated with navigating the food system and using it in order to ensure a regular food intake that is consistent with nutrition recommendations”^33^. This definition of food literacy is inclusive of behavioral and knowledge-based components of literacy. Likewise, nutrition literacy has been defined as “the degree to which individuals have the capacity to obtain, process, and understand nutrition information and skills needed in order to make appropriate nutrition decisions”^34^. This manuscript focuses on nutrition literacy as it relates to facts displayed on FDA compliant food labels. A 2017 review of health literacy as it pertains to nutrition label use and understanding concluded that the “The empirical relationships between health literacy, literacy, numeracy and nutrition label understanding and use have not been well-studied. Additional attention is needed regarding the measurement-related issues identified in the present review”^35^. Accordingly, Nutrition Able has three goals as a project:

1. Develop a novel, accurate, and efficient measurement device for assessing childhood nutrition fact label literacy
2. Increase nutrition fact label literacy through community engagement
3. Assess nutrition fact label literacy in the community

Previous assessment tools proved too lengthy, difficult, or too subjective to serve as a standardized method of studying childhood nutrition facts literacy in a high volume, time constrained classroom setting as is exhibited in this project^36^. Weiss et. al. developed a scientifically validated, publicly available nutrition facts label literacy assessment tool called “The Newest Vital Sign” (NWS)^37^. The device takes less than 3 minutes to complete and contains a nutrition facts label with 6 pertinent questions. NWS would be a sufficient device if

Nutrition Able was carried out in an adult cohort. Studies have been conducted administering NWS to children which confirm poor performance in school-age children^38^. In a letter to the editor, Weiss suggested that the scientific community not focus on developing tools used to assess childhood health literacy^39^. However, the increasing prevalence of childhood obesity and diabetes in San Antonio suggests there is demand for such a tool - as of 2013, the CDC estimated that 32.4% of children in Bexar county ages 10-17 were obese^40^. It is likely that developing appropriate tools to understand childhood food choices can inform clinicians and public health agencies how to better serve their communities.

## Methods

### Study Rationale

Given the disparity in health outcomes between those who live in FDs and those who do not, actionable determinants of health must be identified. Finding gaps in understanding of nutrition labels in FDs is particularly important due to the high prevalence of foods having mandated nutrition facts labels in FDs. As such, NALA is a quick and easy tool which can inform tailored counseling for helping children navigate their food environments.

### Hypotheses

Our study’s primary objective is to determine in a pilot fashion whether NALA is a viable tool for assessing food label literacy. Our study’s primary scientific objective is quantifying differences in nutrition facts label literacy between schools inside or outside of FDs. Secondary objectives include improving these scores, and quantifying the relationship between scores and family history, BMI, and sex. Each explicit hypothesis is listed below.

H1: NALA is a viable tool for measuring nutritional literacy.

H2: Attending school in a FD will be associated with lower NALA pre-survey scores.

H3: Participation in the Nutrition Able program will be associated with improvements NALA survey scores from pre- to post-surveys.

H4: Reporting a family history of obesity and diabetes will be associated with lower NALA pre-survey scores.

H5: A BMI in the overweight or obese ranges (as determined by the CDC growth chart using self-reported height and weight) will be associated with lower NALA pre-survey scores.

H6: There is an association between sex and NALA pre-survey scores.

H7: Attending school in a FD will be associated with higher rates of self-reported family history of obesity and diabetes.

### Study Design and Sampling Method

Our study utilized a cross-sectional design with a novel questionnaire (Figure 1). Participants were asked 7 questions about concepts relevant to a nutrition fact label (and 5 demographic questions. A business card (Figure 2) with a summary of the information in our lecture was dispensed. NALA was administered before giving the lecture about how to read a nutrition label, and was readministered afterwards.

**Figure 2:**
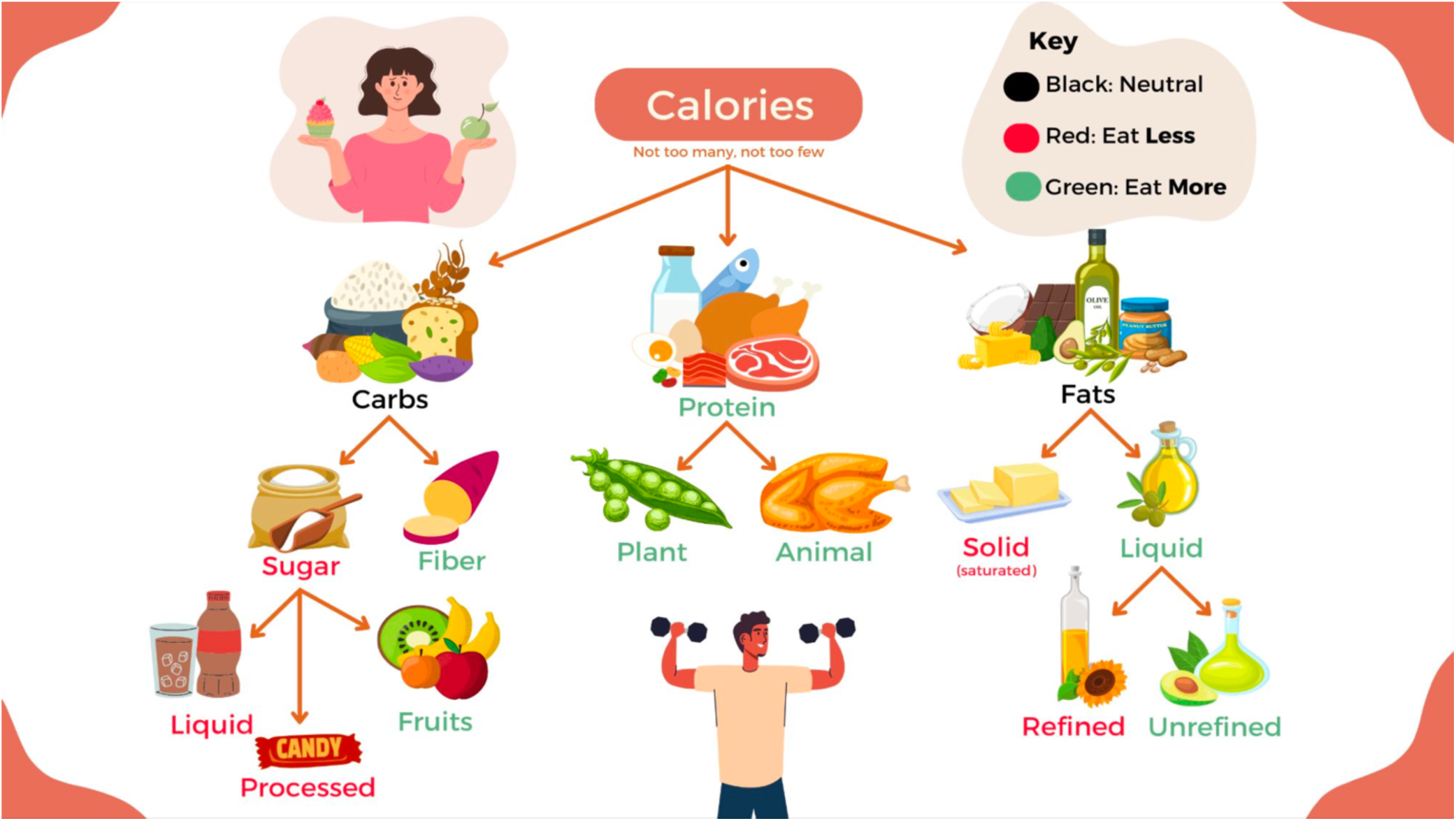

During the first two years that Nutrition Able was funded, our team gave 54 lectures to 12 Texas schools, reaching 1092 students. These talks were given in health, physical education, and biology classes, the vast majority of which were located at schools in FDs. We utilized purposive sampling for convenience, as directed by the school administration. All survey data collected was recorded in REDcap and analyzed for pre/post-score performance comparisons regardless of whether they were fully completed. For demographic analysis, only participants who filled out the optional demographic sections were analyzed. Untouched pre/post-surveys were excluded.

While a total of 1092 students filled out the survey, not all are analyzed for hypotheses 2-7 due to a lack of non-food desert cohort to compare to in the high school age group. This paper analyzes 4 lectures meeting our inclusion/exclusion criteria. These lectures took place in 3 middle schools, looking at 7^th^ and 8^th^ grade students. Two of these schools are in a FD in Bexar County, where the nearest major grocery store, a Walmart Supercenter, was 11.4 and 13.4 miles away, respectively. The third school is in McLennan County and is not in a FD, with multiple nearby grocery stores, the nearest being a Target 1.7 miles away. Our sample consisted of nearly all 7^th^/8^th^-grade students enrolled in the schools, with exceptions including absent students and those who forewent the survey.

The sample of 4 lectures was isolated also because it is free from confounding by school-specific health course curriculum. The schools attended offer health courses in high school at varying grades, making for difficult standardization between schools. None of the students analyzed had received any formal training in nutritional literacy via their school’s curriculum. Furthermore, we have no data on non-food deserts high schools to date, so we are restricted to only comparing the 7^th^ and 8^th^ graders during this analysis. Subsequent analyses will investigate the effects of a FD on high school students’ nutritional literacy.

### Data Collection Process

Surveys (Figure 1) are placed on students’ seats at lectures. The survey consists of two identical pages with an English and Spanish version on the front/back of each page. Students are told the following:

1. The survey is anonymous.
2. The survey is not a test.
3. The survey helps our team assess our project’s success and learn about our community.
4. The survey is optional, but it would be helpful if it was filled out fully.
5. No help will be given.
6. If you do not know an answer or a word, answer to the best of your ability and move on.
7. You will be given ∼4 minutes to complete the survey.

### Variables

- Independent variables are height, weight, age, sex, first-degree family history of obesity and/or diabetes, grade level, and location.
- Dependent variables include overall scores and question-specific correct answer percentages. Overall scores were quantified in our analysis by using the number of correct questions (out of 7) on the questionnaire.

### Data Analysis

Data was handled and analyzed using Microsoft Excel and REDcap. The statistical tests employed include the following:

1. Paired t-tests for analysis of pre/post-scores.
2. One- and two-sided t-tests assuming equal variance for analysis of pre-survey scores by demographic.
3. Chi-squared tests for analysis by demographic if using qualitative data.

To quantify the program’s overall effectiveness, missing surveys were counted as surveys that received a score of 0/7. However, specifically for demographic analyses, surveys missing key information were either excluded or included as an “abstaining” category.

Study data were collected and managed using REDCap electronic data capture tools hosted at University of Texas Health Science Center San Antonio. 1,2 REDCap (Research Electronic Data Capture) is a secure, web-based software platform designed to support data capture for research studies, providing 1) an intuitive interface for validated data capture; 2) audit trails for tracking data manipulation and export procedures; 3) automated export procedures for seamless data downloads to common statistical packages; and 4) procedures for data integration and interoperability with external sources.

## Ethical Considerations

This project is UTHSCSA IRB exempt research under the protocol number 20230587NRR.

## Results

### Baseline Characteristics

Baseline characteristics of the middle school sample (n=235) are shown in Table 2. There were significant differences between groups in all demographics collected except for age. A family history of obesity and diabetes was 27% more likely in the FD (P<0.001). We cross-referenced BMIs with CDC age-specific growth charts to classify each participant into a weight category. There was a significant shift towards greater BMIs in the FD (P<0.05). Some of the analysis by BMI category shown later in this paper is constrained by low sample size in the overweight, obese, and severely obese categories in the NFD. The single NFD participant who self-reported severe obesity by BMI did not answer the questions.

There were significant differences between the proportions of students in 7^th^ vs. 8^th^ grade in the middle school sample (n=235) with the NFD having more 7^th^ graders and the FD having more 8^th^ graders (P<0.001). However, grade level was not a statistically significant predictor of differences in pre/post-survey scores.

Within the middle school sample (n=235), there were significant differences in the proportions of sexes from the FD versus the NFD. As sex was not a significant correlate to pre- or post-survey scores, it is unlikely that the difference in proportion skews the data. In previous abstracts related to this data set^41^, our sampling led us to believe that boys tended to score significantly higher on pre-surveys over a large and robust sample (n=432). In the current analysis, boys indeed scored higher on pre-survey scores. However, girls tended to score higher on post-survey scores. Neither of these trends reached significance.

Previously published analyses showed that boys scored better on pre-surveys with no differences between the sexes in post-surveys (n=432)^41^. The post-survey equivalence had one notable exception: the girls scored significantly worse on the “protein” question in both pre-and post-surveys (n=432)^42^. However, over the entire sample of all students of various grades and locations who have completed NALA, boys do not score significantly better than girls/abstaining participants on pre-survey scores (boys = 3.1 vs. girls = 3.0 vs. abstaining = 2.9; P=0.344). We believe that the lack of statistical significance can be explained by the intentional addition of more females to our presenting team. Over the first 704 surveys, 666 were presented by a male lead presentor with no female present-506 of them were presented by the same duo of male presenters. As the project grew, the Nutrition Able team expanded from one to ten individuals, inclusive of 4 female presenters. With a female presenter, we noted increases in female presurvey scores. In four talks with only female presenters (n=61), girls (n=38) did better than boys (n=23) (3.8 vs. 3.2; P=0.278) on pre-surveys. This suggests that the presence of a same-sex presenter can influence participation at the outset of a discussion about nutrition, and influence the participants’ interpretation of the presentation. Further examination in future papers with larger samples will attempt to quantify this effect with a properly powered analysis.

Nutrition Able was successful at improving nutrition label literacy in middle school students. When comparing pre-/post-surveys (Table 1), mean score increased (2.88 vs. 3.31; P<0.05), median score increased (3 vs. 4) and mode score increased (2 vs. 4). The data show 0 participants scored a perfect 7/7 on the pre-survey, compared to 8 participants on the post-survey (P<0.01). Similarly, 7 participants scored a 6/7 on the presurvey, whereas 24 participants scored a 6/7 on the post-survey (P<0.01). Table 6 shows a pre/post comparison of FD vs. NFD scores, showing significant differences between groups and significant improvements in scores pre/post.

**Table 1.**
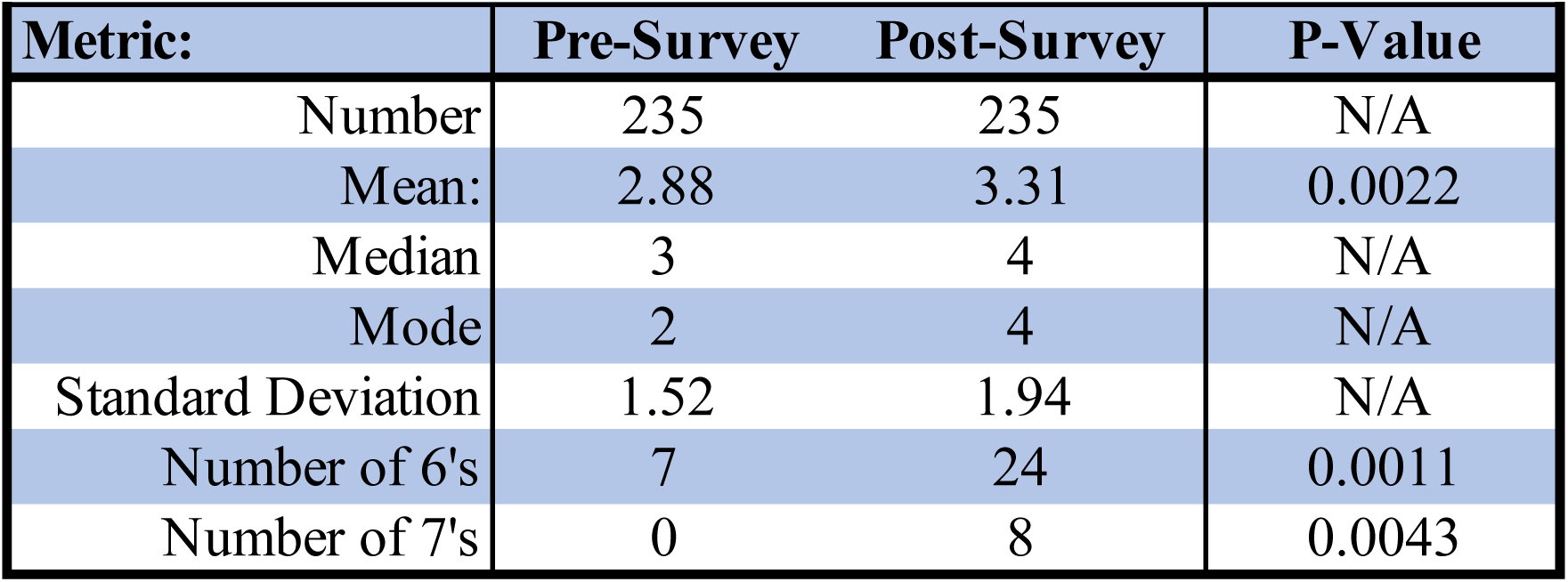
Overall Results (Number of Questions Correct)

**Table 2:**
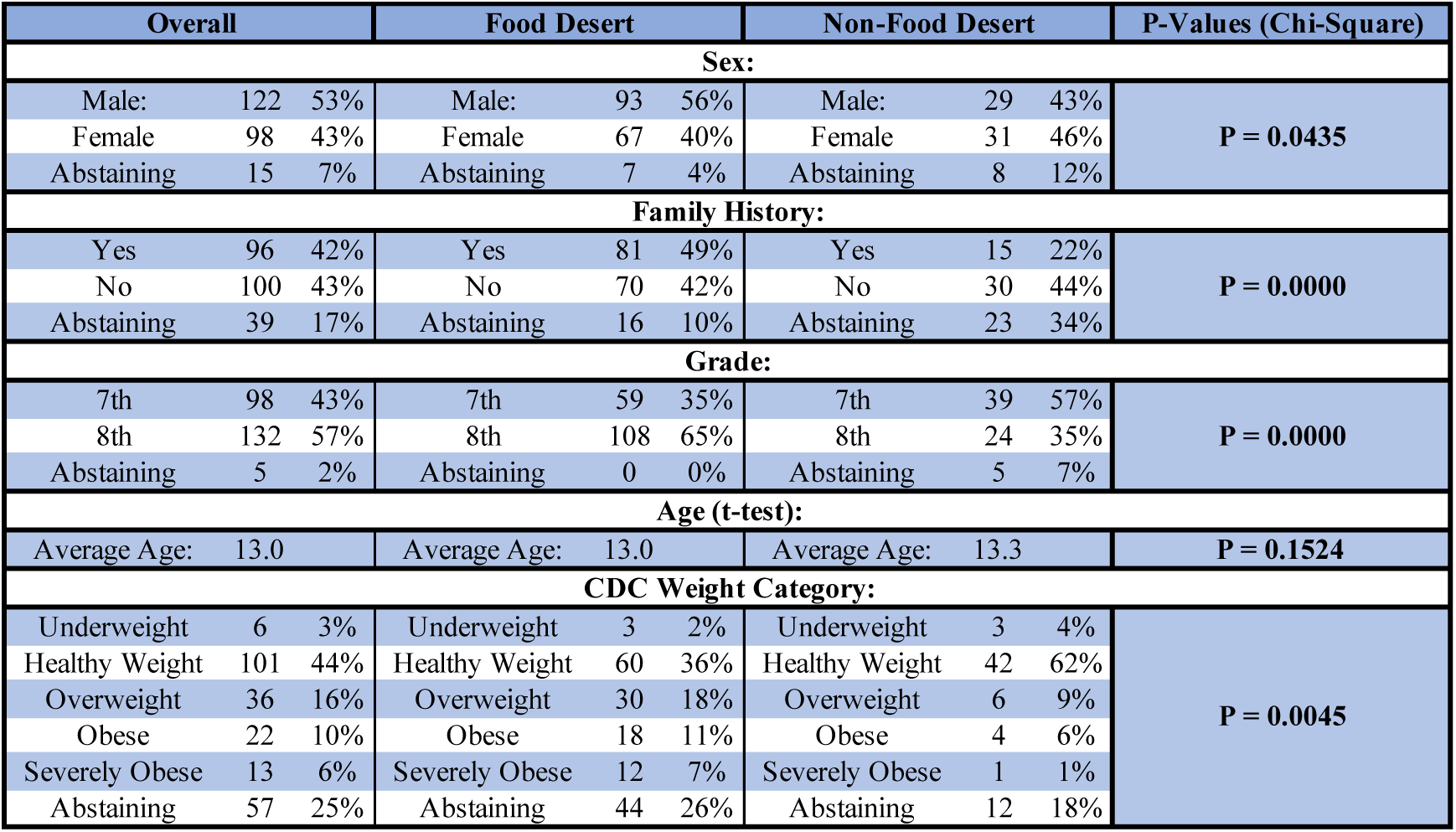
Demographics.

| Table 2: Demographics |  |  |  |  |  |  |  |  |  |
| --- | --- | --- | --- | --- | --- | --- | --- | --- | --- |
| Overall |  |  | Food Desert |  |  | Non-Food Desert |  |  | P-Values (Chi-Square) |
| Sex: |  |  |  |  |  |  |  |  |  |
| Male: | 122 | 53% | Male: | 93 | 56% | Male: | 29 | 43% | P = 0.0435 |
| Female | 98 | 43% | Female | 67 | 40% | Female | 31 | 46% |  |
| Abstaining | 15 | 7% | Abstaining | 7 | 4% | Abstaining | 8 | 12% |  |
| Family History: |  |  |  |  |  |  |  |  |  |
| Yes | 96 | 42% | Yes | 81 | 49% | Yes | 15 | 22% | P = 0.0000 |
| No | 100 | 43% | No | 70 | 42% | No | 30 | 44% |  |
| Abstaining | 39 | 17% | Abstaining | 16 | 10% | Abstaining | 23 | 34% |  |
| Grade: |  |  |  |  |  |  |  |  |  |
| 7th | 98 | 43% | 7th | 59 | 35% | 7th | 39 | 57% | P = 0.0000 |
| 8th | 132 | 57% | 8th | 108 | 65% | 8th | 24 | 35% |  |
| Abstaining | 5 | 2% | Abstaining | 0 | 0% | Abstaining | 5 | 7% |  |
| Age (t-test): |  |  |  |  |  |  |  |  |  |
| Average Age: | 13.0 |  | Average Age: | 13.0 |  | Average Age: | 13.3 |  | P = 0.1524 |
| CDC Weight Category: |  |  |  |  |  |  |  |  |  |
| Underweight | 6 | 3% | Underweight | 3 | 2% | Underweight | 3 | 4% | P = 0.0045 |
| Healthy Weight | 101 | 44% | Healthy Weight | 60 | 36% | Healthy Weight | 42 | 62% |  |
| Overweight | 36 | 16% | Overweight | 30 | 18% | Overweight | 6 | 9% |  |
| Obese | 22 | 10% | Obese | 18 | 11% | Obese | 4 | 6% |  |
| Severely Obese | 13 | 6% | Severely Obese | 12 | 7% | Severely Obese | 1 | 1% |  |
| Abstaining | 57 | 25% | Abstaining | 44 | 26% | Abstaining | 12 | 18% |  |

As seen in tables 3 and 4, Nutrition Able prioritizes time spent teaching participants about the concepts of calories, macronutrients, and calories per serving. The related questions ^(Footnote 1)^ have the largest jumps in pre-/post-survey scores (27.1%; P<0.001, 19.8%; P<0.001, 18.0%; P<0.001). The question which improved the least is “macronutrient content ^(Footnote 2)^”, which starts at a low pre-survey performance relative to the other questions (Pre = 21.5%, Post = 23.6%; P=.5995). Pre-/post-performance on this question did not significantly improve in either group. However, the NFD group had better pre- (42.4% vs. 13.0%; P<0.001) and post-survey scores (42.3% vs. 17.0%; P<0.001) than the FD group. The only other question that did not improve significantly from pre- to post-surveys overall was the “protein” question. When excluding this question, statistical tests of the overall difference between literacy scores in combined, FD, and NFD still reached significance (P<0.001) on both pre- and post-surveys.

**Table 3.**
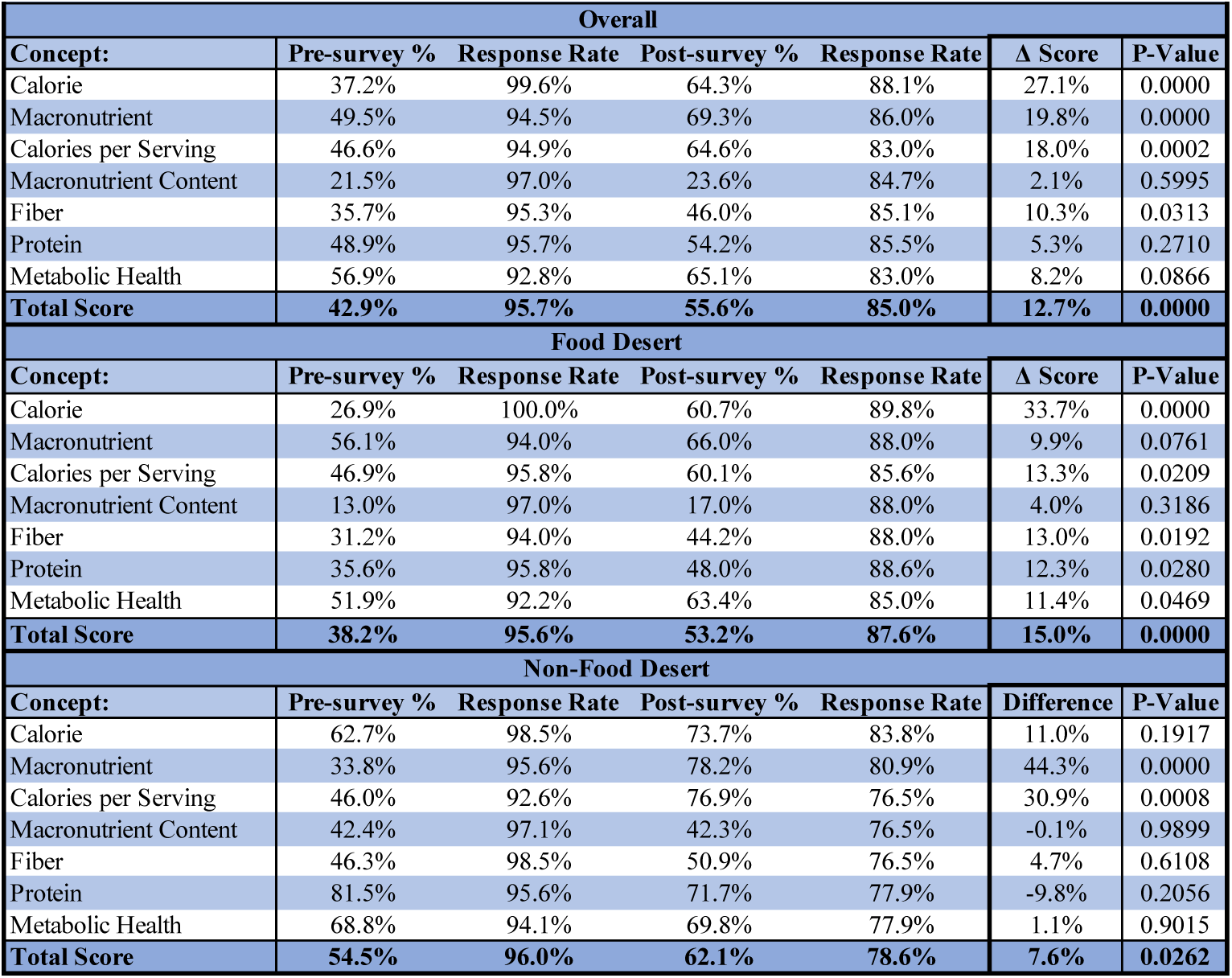
Pre/Post Question by Question Breakdown.

**Table 4.** Food Desert vs. Non Food Desert Question Breakdown.

| Table 4: Food Desert vs. No Food Desert Question Breakdown (Chi-Square Test) |  |  |  |  |
| --- | --- | --- | --- | --- |
| Pre-Survey |  |  |  |  |
| Concept: | Food Desert | Non-Food Desert | Δ Score | P-Value |
| Calorie | 26.9% | 62.7% | 35.7% | 0.0000 |
| Macronutrient | 56.1% | 33.8% | -22.2% | 0.0026 |
| Calories per Serving | 46.9% | 46.0% | -0.8% | 0.5970 |
| Macronutrient Content | 13.0% | 42.4% | 29.5% | 0.0000 |
| Fiber | 31.2% | 46.3% | 15.1% | 0.0313 |
| Protein | 35.6% | 81.5% | 45.9% | 0.0000 |
| Metabolic Health | 51.9% | 68.8% | 16.8% | 0.0225 |
| **Total Score** | **38.2%** | **53.1%** | **14.9%** | **0.0000** |
| Post-Survey |  |  |  |  |
| Concept: | Food Desert | Non-Food Desert | Δ Score | P-Value |
| Calorie | 60.7% | 73.7% | 13.0% | 0.0809 |
| Macronutrient | 66.0% | 78.2% | 12.2% | 0.0944 |
| Calories per Serving | 60.1% | 76.9% | 16.8% | 0.0302 |
| Macronutrient Content | 17.0% | 42.3% | 25.3% | 0.0002 |
| Fiber | 44.2% | 50.9% | 6.7% | 0.3996 |
| Protein | 48.0% | 71.7% | 23.7% | 0.0029 |
| Metabolic Health | 63.4% | 69.8% | 6.4% | 0.4018 |
| **Total Score** | **53.2%** | **62.1%** | **8.9%** | **0.0030** |

In initial analysis, we discovered that the protein question was the only question where there was a significant difference in both pre and post-survey scores when comparing boys to girls (P=0.01). This finding aligns with previous findings in the literature that adolescent females aged 14-18 are the most likely demographic to be consuming protein at a rate below the Estimated Average Requirement^43^. This aligns anecdote suggesting that girls may be less likely to utilize high protein intake and resistance training for fear of drastic changes in bodily appearance.

To investigate whether the initial male/female difference was due to bias, our team incorporated an emphasis on protein intake for women. Osteoporosis is more prevalent in women than men and can be mitigated with protein intake, especially in conjunction with resistance training^44^. A female presenter was included in every talk to determine if a same-sex role model could increase engagement of female participants in hopes that it would resolve the disparity in pre- and post-survey performance. When examining protein question answers with a female presenter (n=286, entirely high school student sample in FD), we noted that pre-survey percentages were not different by sex (male: 65.7%, female: 68.4%, P=0.656). We note a small and insignificant difference in post-survey performance (male: 75.2%, female: 67.4%, P=.183). There was a 1.0% decrease in female pre/post-performance on the protein question (P=0.856) and a 9.5% increase in male pre/post-performance on said question (P=.130). These results indicate an improvement in female performance on the protein pre-survey question with a female presenter.

The starkest difference in any question (n=235) from FD to NFD (excluding the confounded protein question) was the difference in knowledge of a calorie (NFD pre-survey: 62.7%, FD pre-survey 26.9%; P<0.001). Out team values calories as the most important information nutrition facts label for health^22,45^. Nutrition Able successfully decreased the difference in post-survey literacy of calories when looking at NFD vs. FD (60.7% vs. 73.7%; P=0.0809).

In the demographic subgroups analysis (Table 5), only overweight/obese BMI was predictive of score differences. This may be due to small sample sizes, as we had several subgroups which had fewer than 10 surveys total resulting in an underpowered analysis for these groups. When n<5, we qualified numbers in Table 5 with “**”, and when n=0, we qualified numbers in Table 5 with “***”. There were no severely obese participants who completed NALA in the NFD and very few underweight individuals overall. Informed by previous abstracts of a trend^41^, we performed a specific test comparing overweight and obese subjects’ scores with other weight categories’ scores. There was a trend toward better scores and greater score improvements in overweight and obese individuals. In all pre-survey categories (overall, FD, and NFD), overweight and obese patients scored better, and this trend was significant in FDs where there was a larger sample of overweight and obese patients (2.90 vs. 2.43, P=0.025).

**Table 5.**
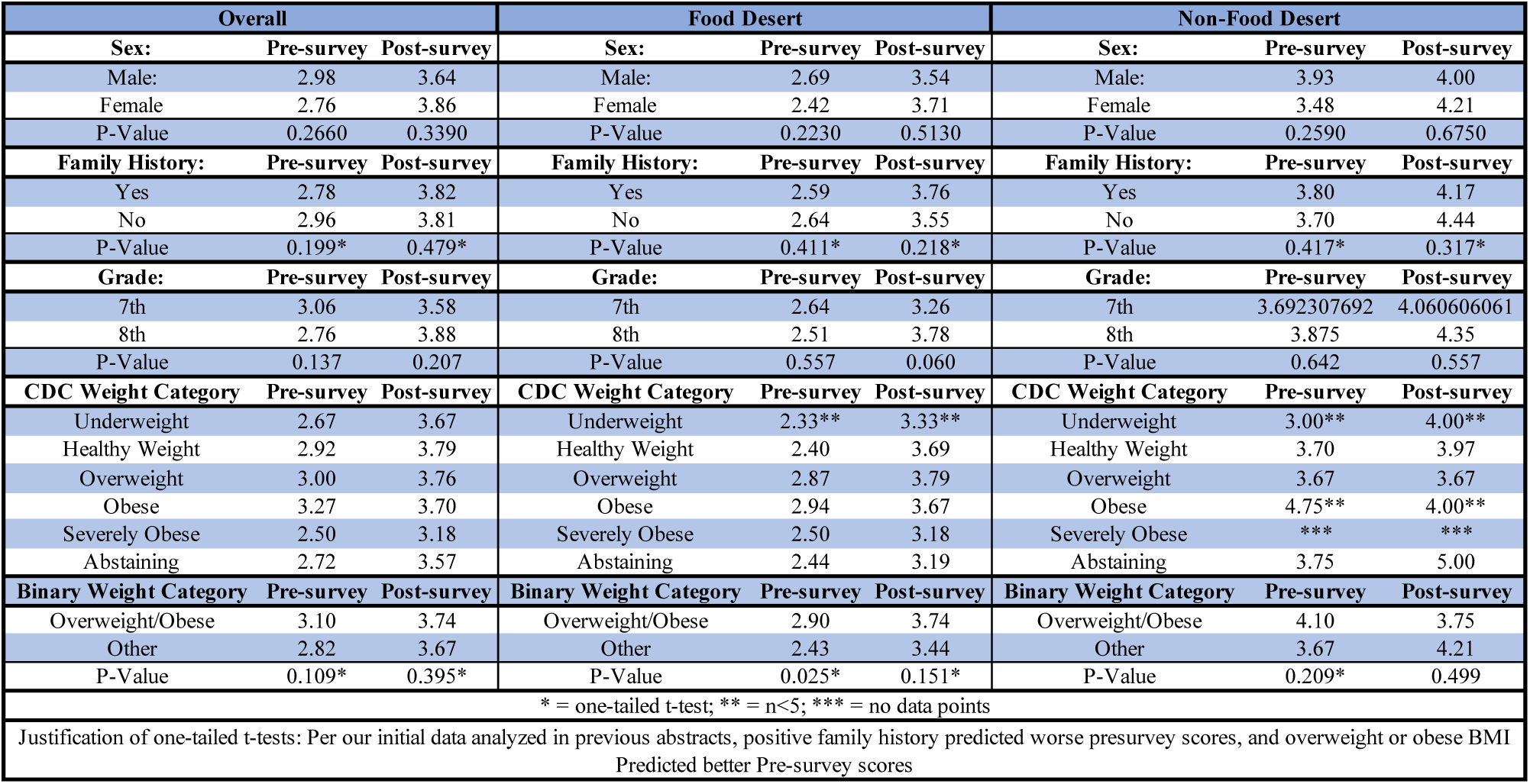
Overall Performance in Number of Questions Correct by Demographic.

**Table 6.**
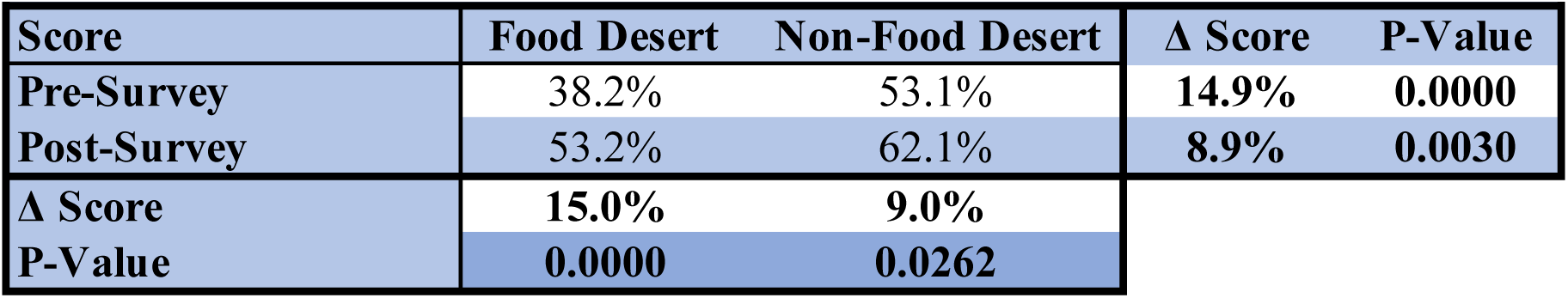
Food Desert vs. Non Food Desert Pre/Post Results.

## Discussion

### Hypothesis 1

Preliminary findings from this pilot study support the hypothesis that the NALA is a viable and responsive tool for measuring nutritional literacy before and after targeted educational interventions. Participants demonstrated statistically significant improvements in post-intervention scores, suggesting that NALA is sensitive to changes in understanding of key nutrition label components, such as serving size calculations, macronutrient interpretation, and the health relevance of fiber and protein. Consistent with prior literature, which suggests that nutrition literacy improves with age and is negatively impacted by limited access to healthy foods^46,31^, our subgroup analyses revealed that older students and those not residing in food deserts scored higher at baseline and showed greater post-intervention gains. While the hypothesis testing below only analyzes 7^th^ and 8^th^ grade students due to the lack of non-food desert samples taken outside of those grades, when including the entire sample of 1092 students, the high school students scored significantly better than the middle school students on both pre-surveys (3.29 vs. 2.48, P<<<0.05) and post-surveys (3.76 vs. 3.26, P<<<0.05). These findings support both the validity of the tool and its capacity to reflect known social determinants of health. Taken together, this pilot supports the use of NALA as a practical, theory-informed assessment instrument to evaluate and guide nutrition education in youth-focused public health efforts.

### Hypothesis 2

Nutrition Able found that students in FDs score worse on pre-survey measures of nutrition label literacy than those not in FDs, supporting our primary hypothesis (n=235, 53.1% vs. 38.2%; P<0.001). This association is corroborated by previous studies evaluating the relationship between food insecurity and health literacy supporting NALA’s use as a tool for evaluating childhood nutrition fact label literacy^32,47^. Per the USDA, a distance of 10 miles from a grocery store warrants classification as a FD-the FD distance was 12 miles, versus 1.7 in the NFD. This classification was by far the most predictive demographic of pre-survey scores. Limitations to the generalizability of this finding include typical confounders to nutritional epidemiology, including but not limited to healthy user bias, socioeconomic differences, self-report, and non-random sampling. Together, these suggest low generalizability of H1. To mitigate these limitations, we adjusted our sample to reduce confounders, as discussed in the methods section. While there were socioeconomic differences between groups, we chose not to collect this demographic within the FD and NFD as the USDA already considers income when classifying FD status.

### Hypothesis 3

Our third hypothesis – that participation in the Nutrition Able program will be associated with improvements NALA survey scores from pre- to post-surveys was analyzed and found to be supported thoroughly in both the FD (n=167, 38.2% vs. 53.2%; P<0.001) and the NFD (n=68, 53.1% vs. 62.1%; P<0.05). In our 7^th^/8^th^ grade sample, inclusive of post-survey non-responders, which were recorded as a 0/7, there was a statistically significant improvement in pre/post-scores from 2.88 to 3.31 (n=235) (P<0.05). The mode score changed from 2/7 to 4/7, and the number of perfect scores went from 0 to 8 (P<0.01). The number of scores of 6/7 went from 7 to 24 (P<0.01). The primary limitation to this analysis is the lack of follow-up over time. This study is cross-sectional in nature and is therefore unable to analyze lasting change in nutrition label literacy.

### Hypothesis 4

Our fourth hypothesis, that reporting a family history of obesity and diabetes would be associated with lower NALA pre-survey scores, was not supported by the data. There was no significant difference in scores when comparing students with a family history to those without^(Footnote 3)^. This analysis is complicated by the lower rates of family history of obesity and diabetes in the NFD population (FD 49% vs. NFD 22%; P<0.001). One study found that family history of hypertension and diabetes are associated with lower health literacy and worse nutritional status. Obesity did not have an association with lower health literacy^48^.

### Hypothesis 5

Our fifth hypothesis, that a BMI in the overweight or obese ranges (as determined by the CDC growth chart using self-reported height and weight) will be associated with lower NALA pre-survey scores, was not supported by the data. An association was discovered in initial analyses showing that overweight and obese BMI’s were actually associated with higher scores, not lower scores (P<0.01)^41,42^. This effect was most pronounced in the FD sample. The lack of significance in the NFD sample is potentially due to an underpowered analysis due to the rarity of overweight and obesity. In the FD, healthy weight individuals scored the worst of any demographic which had n>5. While this hypothesis was not supported, it was noted that there is a gradient in the data sets (both n=235 and the overall n=1092) showing lower literacy scores at the extremes of weight, and lower literacy scores if a participant elects to abstain from reporting their weight. This analysis is subject to the limitation that the BMI’s were calculated from self-reported heights and weights. However, proportions of overweight and obesity in our sample were in line with previous community assessments by San Antonio area governance.

### Hypothesis 6

Our sixth hypothesis, that there is an association between sex and NALA pre-survey scores was not supported by the data. Further analysis and survey collection will seek to discover internal factors which could be influencing survey performance to inform clinicians of how to most effectively improve nutritional literacy in an outpatient youth demographic. For instance, there does appear to be a difference in performance by sex depending on the sex of the presenter. A same-sex presenter was associated with improved performance on both pre- and post-surveys, although this relationship is underpowered for analysis at this time. We recommend programs hoping to set up efficacious food literacy interventions for children to have a diverse core of presenters.

### Hypothesis 7

Our seventh hypothesis, that attending school in a FD will be associated with higher rates of self-reported family history of obesity and diabetes was supported (49% vs. 22%, P<0.001). It is likely that both numbers are significant underrepresentation’s of the true number of students with a family history of obesity and diabetes. Estimations of overweight and obesity rates in San Antonio are around 72%^15^. In 2013, the CDC estimated that 32.4% of children ages 12-17 in Bexar County are overweight or obese. This was reflected in our sample, with 36.0% of students qualifying as overweight or obese. We showed a difference of 27% in prevalence of a family history of obesity and diabetes between NFDs and FDs. There results relied upon self-reported data from children, likely reducing the accuracy of the data. The rate of abstaining from reporting a family history positively or negatively was non-negligible, with 17% of all participants (n=235) abstaining. This rate was much higher in the NFD than in the FD (34% vs. 10%; P<0.001).

## Conclusion

Nutrition Able improved middle and high school students’ scores using a novel and nutrition facts label literacy assessment tool, the Nutrition Able Literacy Assessment (NALA). Future research and clinical assessments should consider use of NALA as a standardized tool for quick assessments in large populations. NALA reliably predicts expected outcomes based on prior data over a large sample size, and has allowed for accurate and consistent assessments of nutrition facts literacy in high school and middle school cohorts.

Attending school in a FD was associated with lower scores, as predicted. The only demographic factor which significantly predicted higher scores was being classified as overweight or obese by the CDC’s growth chart. Severe obesity or underweight was associated with lower literacy scores in both the FD and the NFD. Rates of overweight and obesity self-reported on NALA closely paralleled previous census results, showing that both family history of obesity and diabetes and rates of overweight and obesity were higher in the FD. It is likely that the sex of the presenter effects participant engagement, as evidenced by the increase in scores of the female participants with a female presenter. It is advisable to consider a same sex educator or a diverse presenter team when educating patients and communities about nutrition, although this topic must be explored in detail in future analysis to inform future nutrition literacy interventions.

## Data Availability

All data in the present study are available upon reasonable request to the authors

## Footnotes

1 Questions 1, 2 and 3 in exhibit 1

2 Question 4 in exhibit 1

3 As shown in exhibit 5

