## Supplementary figures and images for "Nutrition Able: A Novel Tool for Improving and Assessing Nutritional Literacy of Middle School Children"

### Supplement 1

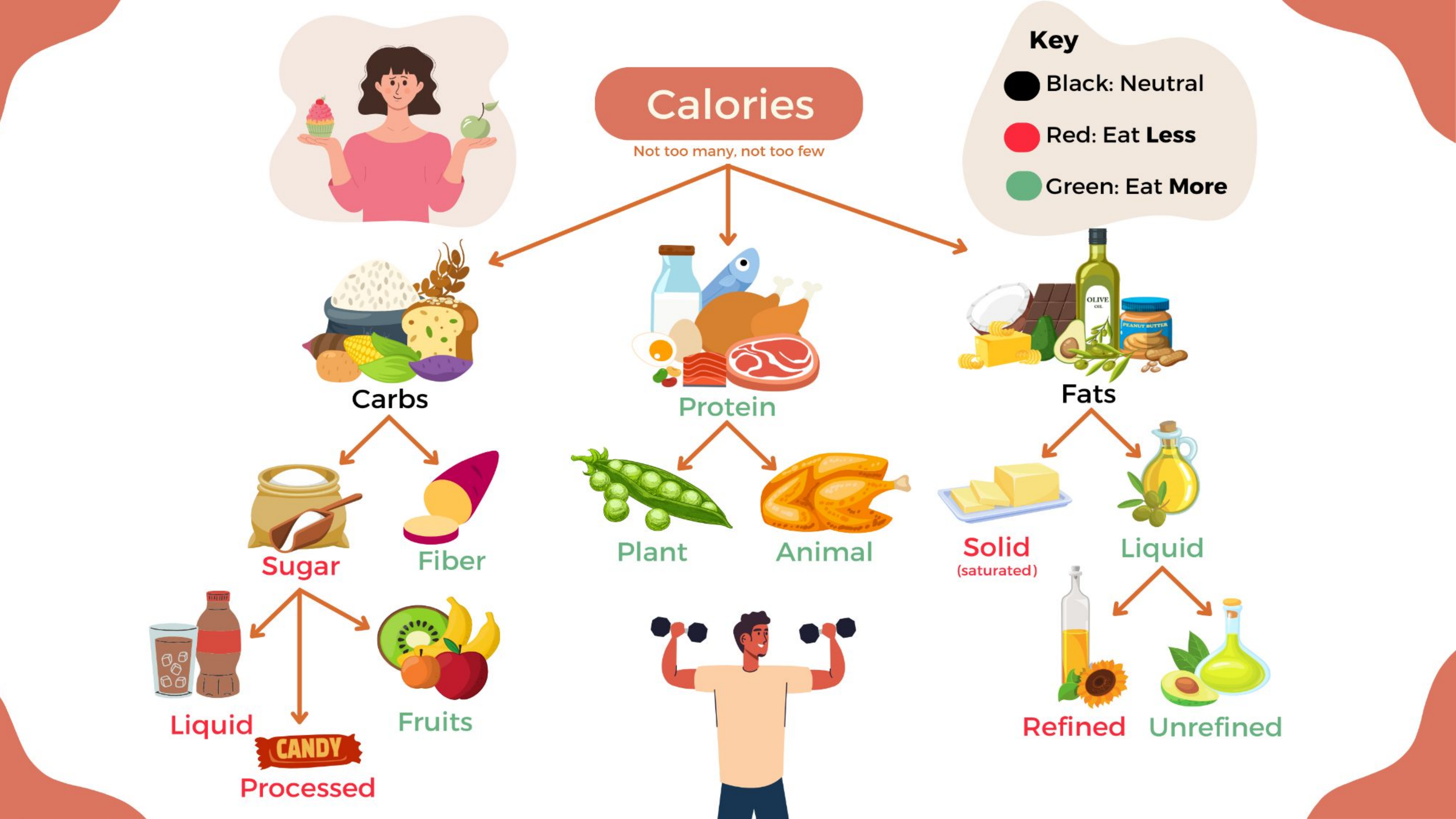
