## Supplement 2 for "Nutrition Able: A Novel Tool for Improving and Assessing Nutritional Literacy of Middle School Children"

UTHSCSA Nutrition Able

### Nutrition Able Literacy Assessment (NALA)

1: What is a calorie?
- A) A unit of weight of a certain food source.
- B) A type of vitamin essential for health.
- C) A unit of energy measurement in food.
- D) A measure of how healthy a food is.

2: What is a macronutrient?
- A) A type of additive found in all processed foods.
- B) A nutrient needed in small amounts for various bodily functions.
- C) A nutrient required in large quantities for energy and growth.
- D) A type of enzyme responsible for digestion.

3: How do you calculate the total calories in a food item with a nutrition label?
- A) (Protein + Fat + Carbs) x 10 = Calories
- B) (Protein x 6) + (Fat x 8) + (Carbohydrates x 5) = Calories
- C) (Weight) / (Macronutrients) = Calories
- D) (Calorie Per Serving) x (Number of Servings) = Calories

4: Which food has the highest carbohydrate content as a percentage?
- A) Milk
- B) Butter
- C) Peanut Butter
- D) Potato
- E) Chicken breast

5: Why is fiber important?
- A) It helps regulate body temperature and causes you to lose weight.
- B) It provides essential vitamins and minerals not found in other foods.
- C) It supports muscle growth and repair by increasing tendon strength.
- D) It maintains bowel health and helps lower blood sugar.

6: Which of the following is true about protein?
- A) It increases muscle mass which helps you stay healthy regardless of weight.
- B) It decreases your need for micronutrients to help overcome nutritional deficits.
- C) It kills fat cells which allows you to be more metabolically healthy.
- D) It is not useful for weight loss promoting diets.

7: What is the most effective home intervention for preventing diabetes/obesity and promoting physical health?
- A) Taking daily multivitamin supplements which increase your body's stores of nutrients.
- B) Increasing the amount of calories you eat by eating more organic foods.
- C) Engaging in regular physical activity, maintaining a balanced diet, and reducing stress.
- D) Using herbal remedies to boost metabolism and overcome the risk of lifestyle factors.

8: Age: _________ Height: _________ Weight: _________Male or Female:________

9: Do you have a first degree relative (parents, siblings, aunts/uncles) with diabetes/obesity?

Yes No

10: What grade are you in?

5th 6th 7th 8th 9th 10th 11th $12^{th}$
